# +Mortality and associated factors among people living with HIV newly initiated into care at Bamenda Regional Hospital : A Retrospective Cohort Study

**DOI:** 10.64898/2026.07.17.26358304

**Authors:** Faustus Ajamah, Jude Tsafack Zefack, Lucas Tanlaka Mengnjo, Onesimus Yongwa, Michael Agbor Ashu, Selouis Fuanyi Nkengfua, Huldah Mbinyui, Alice Ketchaji

**Author notes:** Corresponding author: Faustus Ajamah.

## Abstract

**Introduction:** Globally and in Africa, HIV mortality is reported to have decreased over the years. Cameroon’s national mortality rate was 1.7% in 2023, and there exist regional disparities and underreporting. We assessed the mortality and associated factors among People Living with HIV(PLHIV) initiated into HIV care from January 2021 to December 2023 at the Bamenda regional hospital (BRH)

**Methods:** A retrospective cohort study was conducted from June 2024 to December 2024 at the BRH. Using a non-probabilistic consecutive sampling technique, 331 newly initiated participants on ART were included. Data was collected from medical records using a questionnaire and analyzed using R software version 4.3.1. A multivariable Cox regression model included variables that showed statistical significance and assessed their relationship with mortality while adjusting for potential confounders. Significant predictors of mortality were identified.

**Results:** The mean age of participants was 41.6 ± 11.7 years, and females constituted 57.7%. Opportunistic diseases were present in 19.0% of cases, with tuberculosis (8.2%) being the most common. Key comorbidities included hypertension (8.8%), diabetes (3.9%), and hepatitis B (3.0%). Blood tests showed elevated liver enzymes (ALAT/ASAT: 41.7±23.6 U/L) and high blood sugar (144.4±7.4 mg/dL). In contrast, kidney function (Creatinine: 0.9±0.2 mg/dL) and immune cell counts (Lymphocytes: 1.8±0.7 ×10^3^/μL) appeared normal overall. A cumulative mortality rate of 7.6% over three years (approximately 2.5% per year) was observed. Male sex (AHR=7.83, 95% CI :1.44-42.5, p=0.017), opportunistic diseases (AHR=5.66, 95% CI :1.71-18.7, p=0.004), comorbidities (AHR=6.04, 95% CI :2.02-18.1, p=0.001), Abnormal ALAT/ASAT (p=0.005), and WHO clinical stage III (AHR : 1.1, 95%CI : 1.01 -1.5, p < 0.001), were identified as predictors of mortality. Whereas, BMI and ART adherence showed no significance.

**Conclusion:** Mortality among PLHIV in this study was associated with advanced disease, opportunistic diseases, commorbidities and abnormal laboratory findings. Strengthening early diagnosis, management of opportunistic infections, and monitoring of clinical and laboratory indicators may help reduce mortality in this population.

## 1. Introduction

HIV/AIDS, remains a public health concern with high morbidity and mortality [1]. Since its discovery in 1981, about 78 million individuals have been infected[2]. By 2014, there were an estimated 37 million HIV-positive people globally, with 70% of them living in Sub-Saharan Africa[3]. HIV targets CD4-positive cells in the human body as its host cells, lowering the number and quality of these immune cells and increasing the risk of morbidity and opportunistic infections [2]. A person’--s initial illness occurs two to six weeks after HIV infection because the virus targets cells in the immune system. However, after this initial phase, the virus stays dormant, progressively decreasing the helper T cells in the immune system[4]. Upon reactivation following its latent phase, HIV swiftly attacks T cells, significantly diminishing the immune system’--s efficacy [5]. AIDS is diagnosed when HIV reduces the count of helper T cells to less than 200 cells per microliter of blood, permitting opportunistic infections to occur in the patient[6].

Antiretroviral therapy (ART) is one of the medical advances that has greatly improved the lives of people living with HIV/AIDS (PLHIV), the outcome is reflected in an overall reduction in morbidity and mortality from HIV-related opportunistic infections (OI) as a result of durable suppression of HIV replication and restoration of the body’--s ability to fight against OI[7]. Since 2005, the availability of antiretroviral medication (ART) has expanded; by the end of 2017, an estimated 21.7 million individuals were getting ART globally, and 940,000 people had died from AIDS-related causes [8].

Many patients pass away after beginning therapy, even though the introduction of ART has reduced mortality and morbidity significantly among PLHIV[9]. The literature states that for ART to be effective, patients must understand the significance of ART in their lives, adhere to treatment, and have routine patient follow-up [10]. Patients who are successful on antiretroviral therapy (ART) have an AIDS-related death rate that varies from 4.5% to 17% worldwide[11]. In certain sub-Saharan African populations, there has been evidence of a 17.5% per 100 fatality rate among patients undergoing antiretroviral therapy [12].

In Cameroon, with about 490,484 PLHIV in 2023(2.1% HIV prevalence), the WHO, MoH, and several other NGOs are mobilized to provide HIV care to the Cameroonian population[13]. Despite the introduction of free access to antiretroviral therapy in the country since 2007, the death rate among patients with HIV remains high within the first year of starting ART in some study populations, often due to late diagnosis, advanced disease at presentation, poor adherence, or coexisting opportunistic infections[14].The survival and epidemiology of HIV in Cameroon are influenced by the study population, study locations, and study periods.

Meanwhile, in 2023, Cameroon made progress with regards HIV/AIDS response. Amongst others, the number of new infections which have decreased by 26% compared to 2022, and the number of AIDS-related deaths by 16%. These results are the fruit of different interventions implemented by the stakeholders involved in the national response. However, an HIV mortality rate of 1.7% was recorded in 2023, which remains below the global average of 4.5% reported by the WHO in the same year. Despite the substantial efforts of various stakeholders, discrepancies in mortality rates persist both across and within regions. With a reported HIV prevalence of 3% in 2023, in the North West Region, data on mortality trends and associated factors among newly initiated patients in the Northwest Region, particularly at the Bamenda Regional Hospital which serves as the regional referral center for HIV care, remain scarce.Therefore, this study aimed to determine the mortality rate and identify factors associated with death among people living with HIV who were recently initiated into care at the Bamenda Regional Hospital Treatment Center over a three-year period.

## 2. Methodology

This study employed a retrospective cohort design and was carried out at the Bamenda Regional Hospital Treatment Center. Over a seven-month period, from June to December 2024, Clients files initiated into HIV care between January 2021 and December 2023 were followed retrospectively from the date of ART initiation until death to the end of the observation period. The maximum study period was three years. A non-probability consecutive sampling approach was used to select files of patients who had been initiated on ART between January 2021 and December 2023. Follow-up time was defined as the period from ART initiation to death or last clinic visit recorded before December 2023. Data were extracted using a pretested structured questionnaire, organized into four sections: Section A captured patient identification details; Section B recorded socio-demographic characteristics; Section C focused on HIV clinical data; and Section D covered biological parameters. Of the 356 patient files initially recruited, 12 were classified as lost to follow-up (LTFU) and 13 were transferred out to other health facilities, leaving the remaining 331 records for final analysis. This systematic process ensured comprehensive data collection for analysis.

### 2.1 Statistical analysis

Data from the questionnaires was input into CSPro version 7.5 software and exported to MS Excel for cleaning. Finally, it was exported to the R software version 4.3.1 for analysis. Descriptive statistics were used to summarize data. Continuous variables were expressed as mean ± standard deviation for normally distributed variables, median (interquartile range) for skewed variables, and proportions and frequencies for categorical variables. For inferential analysis, bivariate Cox proportional hazards regression was performed to identify predictors of mortality. Variables with a p-value < 0.20 in the bivariate analysis were entered into the multivariable Cox regression model to examine the independent association between multiple variables and mortality while controlling for potential confounders. This approach enabled the identification of the most significant predictors of mortality and provided insight into how different factors interact to affect mortality rates.

### 2.2 Declarations

- **Ethical approval** Ethical approval was obtained from the Institutional Review Board of the Catholic University of Central Africa (Ref: 2024/02082/CEIRSH/ESS/MSP). All procedures involving human participants were conducted in accordance with relevant institutional guidelines and regulations and in line with the principles of the Declaration of Helsinki.
- **Consent to participate** The requirement for informed consent was waived by the same Institutional Review Board due to the retrospective nature of the study and the use of anonymized routine program data.
- **Consent to publish** Not applicable, as the manuscript does not contain any individual person’--s identifiable data.

## 3. Results

### 3.1 Sociodemographic profile of study sample

In this study, the mean age was 41.6 ± 11.7 years, with the majority aged 40–50 (32.9%). Females constituted a larger proportion (57.7%), and over half were in unskilled occupations (54.7%). Most participants attained primary (40.2%) education, and 45.3% were single. Additionally, two-thirds resided within 10 km of the hospital.

**Table 1:** Sociodemographic Characteristics of the study sample.

| Variables | Category | Frequency (Percentage) |
| --- | --- | --- |
| Age range (years) | 19–29 | 55 (16.6%) |
|  | 30–39 | 93 (28.1%) |
|  | 40–49 | 109 (32.9%) |
|  | 50–59 | 48 (14.5%) |
|  | 60–77 | 26 (7.85%) |
| Sex | Female | 191 (57.7%) |
|  | Male | 140 (42.3%) |
| Age | Mean ± SD | 41.6 ± 11.7 |
| Occupation | Skilled | 150 (45.3%) |
|  | Unskilled | 181 (54.7%) |
| Level of education | No education | 19 (5.74%) |
|  | Primary | 133 (40.2%) |
|  | Secondary | 126 (38.1%) |
|  | University | 53 (16.0%) |
| Marital status | Single | 150 (45.3%) |
|  | Divorced | 14 (4.23%) |
|  | Married | 128 (38.7%) |
|  | Widow/Widower | 39 (11.8%) |
| Residence | <10 km | 220 (66.5%) |
|  | 10–20 km | 78 (23.6%) |
|  | >20 km | 33 (9.97%) |

### 3.2 Clinical profile of study participants

The majority of participants were HIV-1 positive (98.5%), with a 3 year cummulative mortality rate of 7.6% observed during the study period. Most patients (91.5%) reported good ART adherence. Meanwhile, Opportunistic disease, comorbidities, and poor adherence were more frequent among those who died, while most participants (96.7%) were in WHO Stage 1 at the time of initiation.

**Table 2:**
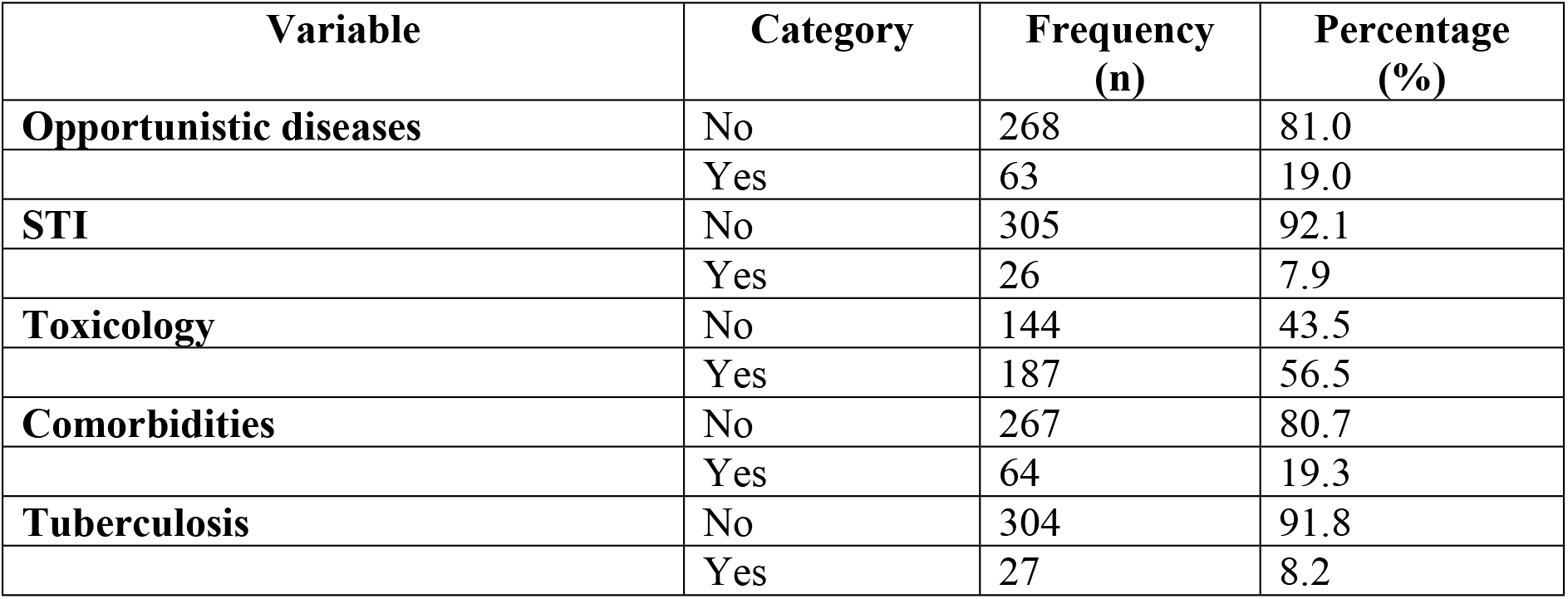
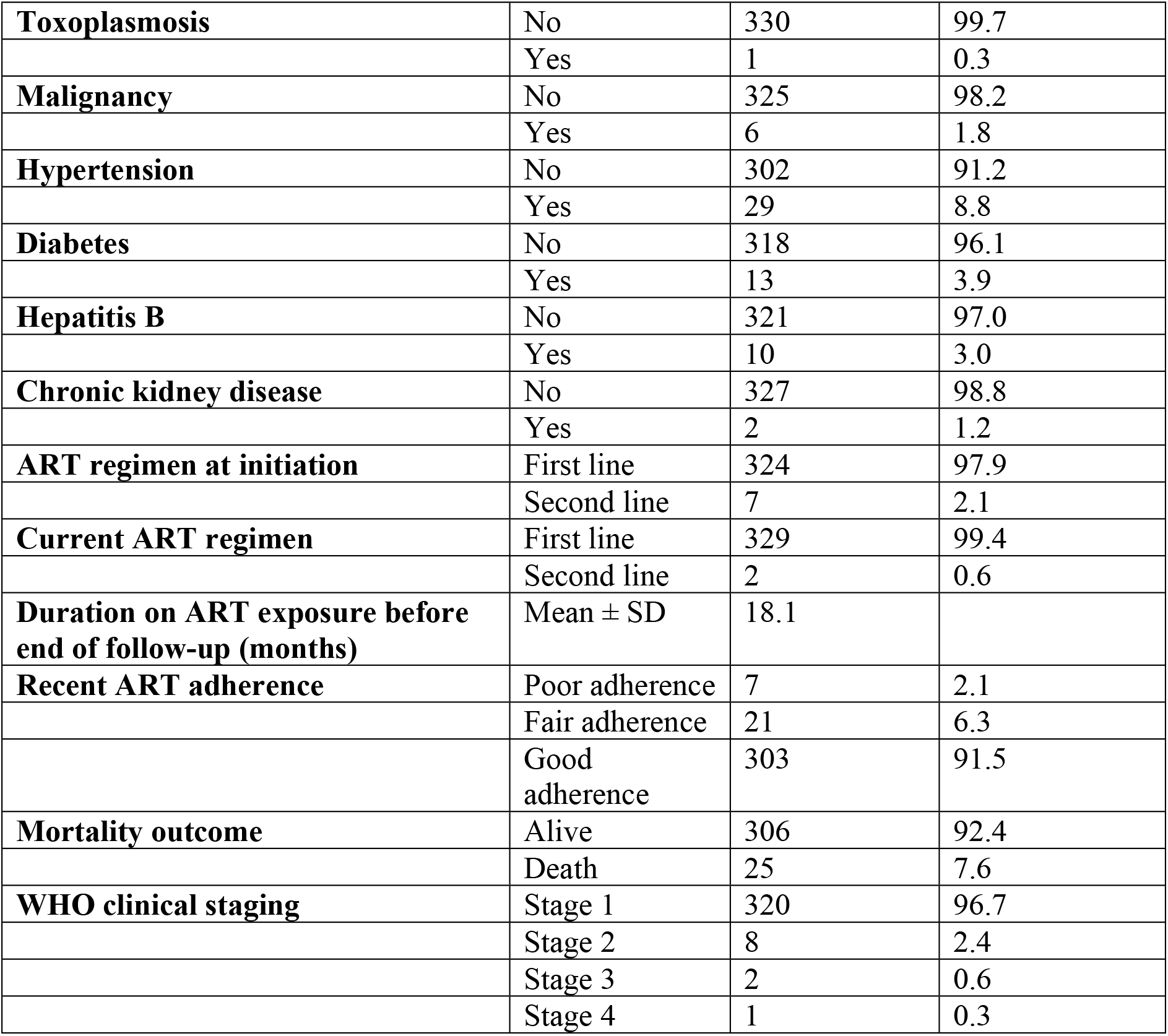
Clinical characteristics of study participants.

| Variable | Category | Frequency (n) | Percentage (%) |
| --- | --- | --- | --- |
| Opportunistic diseases | No | 268 | 81.0 |
|  | Yes | 63 | 19.0 |
| STI | No | 305 | 92.1 |
|  | Yes | 26 | 7.9 |
| Toxicology | No | 144 | 43.5 |
|  | Yes | 187 | 56.5 |
| Comorbidities | No | 267 | 80.7 |
|  | Yes | 64 | 19.3 |
| Tuberculosis | No | 304 | 91.8 |
|  | Yes | 27 | 8.2 |
| **Toxoplasmosis** | No | 330 | 99.7 |
|  | Yes | 1 | 0.3 |
| **Malignancy** | No | 325 | 98.2 |
|  | Yes | 6 | 1.8 |
| **Hypertension** | No | 302 | 91.2 |
|  | Yes | 29 | 8.8 |
| **Diabetes** | No | 318 | 96.1 |
|  | Yes | 13 | 3.9 |
| **Hepatitis B** | No | 321 | 97.0 |
|  | Yes | 10 | 3.0 |
| **Chronic kidney disease** | No | 327 | 98.8 |
|  | Yes | 2 | 1.2 |
| **ART regimen at initiation** | First line | 324 | 97.9 |
|  | Second line | 7 | 2.1 |
| **Current ART regimen** | First line | 329 | 99.4 |
|  | Second line | 2 | 0.6 |
| **Duration on ART exposure before end of follow-up (months)** | Mean ± SD | 18.1 | |
| **Recent ART adherence** | Poor adherence | 7 | 2.1 |
|  | Fair adherence | 21 | 6.3 |
|  | Good adherence | 303 | 91.5 |
| **Mortality outcome** | Alive | 306 | 92.4 |
|  | Death | 25 | 7.6 |
| **WHO clinical staging** | Stage 1 | 320 | 96.7 |
|  | Stage 2 | 8 | 2.4 |
|  | Stage 3 | 2 | 0.6 |
|  | Stage 4 | 1 | 0.3 |

### 3.3 Biological profile of study participants

The mean ALAT/ASAT level was 41.7 ± 23.6 U/L, while mean creatinine was 0.9 ± 0.2 mg/dL. Average glycemia stood at 144.4 ± 7.4 mg/dL, and mean hemoglobin was relatively low at 11.1 ± 1.1 g/dL. Lymphocyte and platelet counts were consistent across participants, with minimal variation noted.

**Tableau 3:** Description of the biological characteristics of the study sample.

| **Variable** | **Mean ± SD** | **Unit** |
| --- | --- | --- |
| ALAT/ASAT | $41.7 \pm 23.6$ | U/L |
| Creatininemia | $0.9 \pm 0.2$ | mg/dL |
| Glycemia | $144.4 \pm 7.4$ | mg/dL |
| Lymphocytes | $1.8 \pm 0.7$ | $\times 10^3/\mu\text{L}$ |
| Platelets | $204.0 \pm 64.2$ | $\times 10^9/\text{L}$ |
| Hemoglobin | $11.1 \pm 1.1$ | g/dL |

### 3.4 Bivariate analysis of study variables with outcome variable

Mortality was higher among males (14.3%) compared to females (2.6%), with male sex significantly associated with death (HR: 5.66; p < 0.001). Older age groups showed increasing hazard ratios, though not statistically significant. Unskilled workers and those with no formal education had higher death proportions, but these associations were not significant. No significant relationship was found between mortality and marital status or distance from the hospital.

**Table 4:** Bivariate analysis of HIV mortality and sociodemographic characteristics.

| Variables | Outcome |  | HR | 95% CI | p-value |
| --- | --- | --- | --- | --- | --- |
|  | Death<br>n = 25 | Alive<br>n = 306 |  |  |  |
| **Age range** |  |  |  |  |  |
| [20-30[ | 1 (1.82%) | 54 (98.2%) | — | — |  |
| [30-40[ | 8 (8.60%) | 85 (91.4%) | 4.53 | 0.57, 36.2 | 0.154 |
| [40-50[ | 11 (10.1%) | 98 (89.9%) | 5.93 | 0.76, 45.9 | 0.089 |
| [50-60[ | 1 (2.08%) | 47 (97.9%) | 1.18 | 0.07, 18.9 | 0.907 |
| [60,77] | 4 (15.4%) | 22 (84.6%) | 8.38 | 0.94, 75.0 | **0.057** |
| **Age** |  |  |  |  | 0.345 |
| Mean ± SD | 43.7 ± 10.8 | 41.4 ± 11.7 | | | |
| **Sex** |  |  |  |  |  |
| Female | 5 (2.62%) | 186 (97.4%) | — | — |  |
| Male | 20 (14.3%) | 120 (85.7%) | 5.66 | 2.13, 15.1 | **<0.001** |
| **Occupation** |  |  |  |  |  |
| Skilled | 8 (5.33%) | 142 (94.7%) | — | — |  |
| Unskilled | 17 (9.39%) | 164 (90.6%) | 1.78 | 0.77, 4.13 | 0.177 |
| **Level of education** |  |  |  |  |  |
| No education | 3 (15.8%) | 16 (84.2%) | — | — |  |
| Primary | 11 (8.27%) | 122 (91.7%) | 0.45 | 0.13, 1.62 | 0.221 |
| Secondary | 8 (6.35%) | 118 (93.7%) | 0.33 | 0.09, 1.25 | 0.103 |
| University | 3 (5.66%) | 50 (94.3%) | 0.28 | 0.06, 1.41 | 0.123 |
| **Marital status** |  |  |  |  |  |
| Single | 11 (7.33%) | 139 (92.7%) | — | — |  |
| Divorce | 1 (7.14%) | 13 (92.9%) | 1.07 | 0.14, 8.33 | 0.945 |
| Married | 11 (8.59%) | 117 (91.4%) | 1.19 | 0.52, 2.75 | 0.683 |
| Widow/widower | 2 (5.13%) | 37 (94.9%) | 0.74 | 0.16, 3.36 | 0.701 |
| **Residence** |  |  |  |  |  |
| < 10 km | 18 (8.18%) | 202 (91.8%) | — | — |  |
| 10-20km | 5 (6.41%) | 73 (93.6%) | 0.74 | 0.28, 2.00 | 0.554 |
| >20km | 2 (6.06%) | 31 (93.9%) | 0.73 | 0.17, 3.16 | 0.677 |

### 3.5 Bivariate analysis of HIV mortality and clinical characteristics

Presence of opportunistic infections (HR: 7.13; *p* < 0.001), malignancies (HR: 28.9; *p* < 0.001), comorbidities (HR: 4.92; *p* < 0.001), and advanced WHO clinical stages at ART initiation (Stage 4: HR: 6.68; *p* < 0.001) were strongly associated with increased mortality. Lower BMI and lack of viral suppression (HR: 5.68; *p* < 0.001) also significantly predicted death. Poor ART adherence had the highest risk (HR: 15.5; *p* < 0.001). HIV type, candidiasis, STIs, and ART regimen were not significantly associated with mortality.

**Table 5:** Bivariate analysis of HIV mortality and clinical characteristics.

| Variable | Category | Death n (%) | Alive n (%) | HR | 95% CI | p-value |
| --- | --- | --- | --- | --- | --- | --- |
| **Opportunistic infection** | No | 10 (3.7) | 259 (96.3) | Ref | — | — |
|  | Yes | 15 (24.2) | 47 (75.8) | 7.13 | 3.20–15.9 | **<0.001** |
| **STI** | No | 24 (7.9) | 281 (92.1) | Ref | — | — |
|  | Yes | 1 (3.9) | 25 (96.1) | 0.43 | 0.06–3.18 | 0.407 |
| **Tuberculosis** | No | 21 (6.9) | 283 (93.1) | Ref | — | — |
|  | Yes | 4 (14.8) | 23 (85.2) | 2.13 | 0.73–6.19 | 0.167 |
| **Malignancy** | No | 20 (6.2) | 305 (93.8) | Ref | — | — |
|  | Yes | 5 (83.3) | 1 (16.7) | 28.9 | 10.4–80.4 | **<0.001** |
| **Toxicology** | No | 8 (5.6) | 136 (94.4) | Ref | — | — |
|  | Yes | 17 (9.1) | 170 (90.9) | 1.69 | 0.73–3.92 | 0.220 |
| **BMI (kg/m<sup>2</sup>)** | Mean ± SD | 21.1 ± 2.4 | 23.5 ± 3.7 | — | — | **0.004** |
| **BMI category** | 18.5–24.9 | 19 (11.1) | 152 (88.9) | Ref | — | — |
|  | 25–29.9 | 1 (1.6) | 62 (98.4) | 0.13 | 0.02–0.98 | **0.048** |
| **Comorbidities** | No | 13 (4.8) | 257 (95.2) | Ref | — | — |
|  | Yes | 12 (19.7) | 49 (80.3) | 4.92 | 2.24–10.8 | **<0.001** |
| **Hypertension** | No | 21 (7.0) | 281 (93.0) | Ref | — | — |
|  | Yes | 4 (13.8) | 25 (86.2) | 2.19 | 0.75–6.39 | 0.151 |
| **WHO clinical stage** | Stage 1 | 17 (5.3) | 303 (94.7) | Ref | — | — |
|  | Stage 2 | 6 (75.0) | 2 (25.0) | 20.4 | 8.00–51.8 | **<0.001** |
|  | Stage 3 | 1 (50.0) | 1 (50.0) | 14.8 | 1.96–111 | **0.009** |
| **ART adherence** | Good | 15 (5.0) | 288 (95.0) | Ref | — | — |
|  | Fair | 5 (23.8) | 16 (76.2) | 6.86 | 2.47–19.0 | **<0.001** |
|  | Poor | 5 (71.4) | 2 (28.6) | 15.5 | 5.60–42.7 | **<0.001** |

### 3.6 Link between Biological characteristics and Mortality outcome

Elevated liver enzymes (ALAT/ASAT) were significantly associated with increased mortality (*p* < 0.001), suggesting hepatic involvement as a potential risk factor. Other biological parameters including creatinine, glycemia, lymphocyte count, platelet count, hemoglobin level, and blood count did not show statistically significant associations with mortality.

**Table 6:** Link between Biological characteristics and outcome variable.

| Variable | Death (n = 25) | Alive (n = 306) | p-value |
| --- | --- | --- | --- |
| **ALAT/ASAT (U/L)** | 42 (25–78) | 27 (18–40) | **<0.001** |
| **Creatininemia (mg/dL)** | 0.90 ± 0.20 | 0.89 ± 0.21 | 0.78 |
| **Glycemia (mg/dL)** | 146.2 ± 18.6 | 144.1 ± 21.3 | 0.64 |
| **Hemoglobin (g/dL)** | 10.9 ± 1.2 | 11.1 ± 1.1 | 0.36 |
| **Lymphocytes ($\times 10^3/\mu\text{L}$)** | 1.7 ± 0.6 | 1.8 ± 0.7 | 0.48 |
| **Platelets ($\times 10^9/\text{L}$)** | 198 ± 61 | 204 ± 64 | 0.62 |

### 3.7 Predictors of mortality among the study population

Male sex (AHR = 7.83, *p* = 0.017), opportunistic infections (AHR = 5.66, *p* = 0.004), comorbidities (AHR = 6.04, *p* = 0.001), and WHO Stage 3 (AHR = 1.10, *p* < 0.001) were significantly associated with higher mortality. Poor ART adherence and BMI showed no significant association.

**Table 7:** Multivariate Cox-Regression Analysis of the study population.

| Variable | Category | AHR | 95% CI | p-value |
| --- | --- | --- | --- | --- |
| **Sex** | Female | Ref | — | — |
|  | Male | 7.83 | 1.44–42.5 | **0.017** |
| **Opportunistic infection** | No | Ref | — | — |
|  | Yes | 5.66 | 1.71–18.7 | **0.004** |
| **WHO clinical stage** | Stage 1 | Ref | — | — |
|  | Stage 2 | 4.66 | 0.89–24.4 | 0.069 |
|  | Stage 3 | 1.10 | 1.01–1.50 | **<0.001** |
| **BMI range** | 18.5–24.9 | Ref | — | — |
|  | 25–29.9 | 0.32 | 0.04–2.67 | 0.293 |
| **ALAT/ASAT (U/L)** |  | 1.03 | 1.01–1.05 | **0.005** |
| **ART adherence** | Good adherence | Ref | — | — |
|  | Poor adherence | 2.88 | 0.48–17.2 | 0.245 |
|  | Fair adherence | 2.83 | 0.53–15.1 | 0.225 |
| **Comorbidities** | No | Ref | — | — |
|  | Yes | 6.04 | 2.02–18.1 | **0.001** |

## 4. Discussion

This study aimed to determine the mortality rate, identify factors associated with mortality following initiation of care among PLHIV attending the BRH treatment center. Male sex, Opportunistic diseases, advanced WHO clinical staging Stage III, abnormal liver enzymes, and comorbidity were significant predictors of mortality after initiation of care. Accounting for these factors may help in refining the prescription of ART and improving the outcome of care among PLHIV attending the BRH treatment center. The mean age in this study was 41.59 ± 11.7 years, with the 40–50 age group (32%) being the most affected. These findings align with Owashi et al. (2024) in Uganda, who reported a mean age of 39 years[15]. This consistency supports existing literature indicating that the disease predominantly affects younger adults. More than half (57%) of the study participants were women. This matches national(Cameroon) HIV distribution data, where most people on HIV care are females(3.4% in females and 1.9% in male**s**)[16].

We examined the mortality rate and correlated socio-demographic, clinical, and biological factors, and observed a mortality rate of 7.6% per 3-year (equivalent to 2.5% annually). These findings indicate significant progress in targeted interventions addressing factors contributing to mortality within this population compared to the 20.2% per year follow-up rate reported by Sieleunou et al. in Cameroon’--s Far North region[17]. This is likely attributable to improved ART access and enhanced early diagnosis. It could also be speculated that this disparity between this previous study and our findings reflects differences in healthcare infrastructure between our referral center and rural treatment facilities, where resource limitations often constrain service provision to basic care. The disparity is further contextualized by Poka-

Mayap et al(2013), who reported a rate of 16.3% per year of follow-up in a referral hospital in Yaounde[18]. Their intermediate rate, which falls between rural figures and ours, might be because their study took place during the early phase of ART scale-up (2007–2011), the time when treatment protocols were still being developed and hadn’--t yet been fully standardized. However, this rate is slightly lower than the 3.95 per 100 person-years reported by Kyaw et al., though both fall within the range of improved outcomes compared to pre-ART era studies[19].

Among the sociodemographic factors associated with mortality, male participants(80%) demonstrated a significantly higher risk of death compared to females (AHR: 7.88; 95% CI: 1.44–42.5; p < 0.017) following care initiation. This sex disparity reflects existing literature, though the magnitude varies across settings: Kyaw et al. reported a more modest male-associated risk (AHR: 1.2; 95% CI: 1.03–1.39), while Cambodian data showed an intermediate hazard (AHR: 1.75; 95% CI: 1.04–3.0)[20]. This pattern may reflect delayed HIV care-seeking behavior among men. Notably, Mbuagbaw et al. (2016) reported a higher proportion of female deaths (54.4%)[21]. This finding contrasts with our results and underscores the need for sex-specific interventions tailored to local epidemiological contexts.

Our study reveals important clinical associations with mortality among PLHIV attending the BRH. The finding that opportunistic infections were associated with a fivefold increase in mortality risk (AHR: 5.66; 95% CI: 1.71-18.7) echoes findings from similar settings in sub-Saharan Africa, such as that of Chimbetete et al. in Zimbabwe[22]. However, our effect size exceeds theirs, possibly reflecting diagnostic delays or limited access to prophylactic regimens in our context. Our findings also showed that late presentation for ART at WHO clinical Stage III was independently associated with an increased hazard of mortality (AHR = 1.10), suggesting that more advanced disease at presentation may adversely affect survival. Although the magnitude of this association was modest, it is consistent with evidence indicating that delayed initiation of HIV care is associated with poorer clinical outcomes. Comorbidities significantly elevated mortality risk (AHR: 6.04 ). These findings align with Nigerian studies and underscore the need for integrated HIV/NCD care[23,24].

Poor viral suppression may reflect adherence challenges or drug resistance, while comorbidities like hypertension or diabetes likely compound mortality due to fragmented care systems. However, the non-significant association between ART adherence and mortality (AHR: 2.88 for poor adherence, p = 0.245) contrasts with the literature and may reflect the adherence measurement limitations[25]. Similarly, BMI showed no significant effect, diverging from Western cohorts where obesity is protective[26]. In this setting, undernutrition, not overweight, may drive outcomes, ratifying the well-established link between wasting and HIV disease progression, emphasizing the role of food insecurity in HIV progression. Furthermore, biological markers in except of ALAT/ASAT, reveal no significant association with mortality. These findings, however, contrast with several other studies, including that of Tadesse et al. in Northern Ethiopia, who reported that both hemoglobin levels and viral load were independently associated with mortality among PLHIV[27]. ALAT/ASAT(p<0.005), revealed a significant associations with WHO clinical stage mortality that complement the clinical picture emerging from our findings, suggesting hepatic involvement may represent an underrecognized contributor to mortality in our cohort. This association likely reflects multiple pathological processes, including drug-induced liver injury from antiretroviral therapy, concurrent viral hepatitis coinfection, or systemic inflammatory responses to advanced HIV disease.

### 4.1 Study Limitations

A key limitation of this study stems from its retrospective design, which resulted in the exclusion of incomplete patient records, with vital missing data such as missing CD4 counts. Additionally, Selection bias might have been introduced since records with incomplete information that were lost for some patients were excluded. On the other hand, participants who were lost to follow-up or transferred were not included in our study, which might have underestimated the mortality rate of the cohort. However, the retrospective cohort study design effectively identified key predictors of mortality. These findings should inform both clinical practice and programmatic decisions as we work to optimize HIV care at the BRH treatment center and similar resource-limited settings. Future research should also explore cultural and psychosocial predictors of HIV mortality, particularly given the ongoing sociopolitical challenges in the North West Region, which may influence patient outcomes and engagement with care.

### 4.2 Conclusion

This study found a cumulative mortality rate of 7.6% among PLHIV initiating ART at Bamenda Regional Hospital over three years. Male sex, opportunistic infections, comorbidities, advanced WHO clinical stage, and elevated liver enzymes were independently associated with mortality. Strengthening early HIV diagnosis, timely initiation of ART, and comprehensive management of opportunistic infections and comorbidities may significantly improve survival outcomes among PLHIV in this setting.

## Data Availability

Data is available to the corresponding author upon reasonable request

## Authorship contributions

FA, and AK contributed to the conception and design of the study. FA, LTM, and HM were responsible for data collection. FA, and JTZ performed data analysis and interpretation. MAA and OY contributed to interpretation of results and provided methodological support. FA drafted the manuscript. AK, JTZ, LTM, OY, MAA, SFN, and HM critically revised the manuscript for important intellectual content.

## Data Availability

The datasets used and/or analysed during the current study are available from the corresponding author on reasonable request.

## References

1. Workie KL, Birhan TY, Angaw DA. Predictors of mortality rate among adult HIV-positive patients on antiretroviral therapy in Metema Hospital, Northwest Ethiopia: a retrospective follow-up study. AIDS Research and Therapy. 2021 May 5;18(1):27.

2. Gunda DW, Nkandala I, Kilonzo SB, Kilangi BB, Mpondo BC. Prevalence and Risk Factors of Mortality among Adult HIV Patients Initiating ART in Rural Setting of HIV Care and Treatment Services in North Western Tanzania: A Retrospective Cohort Study. Journal of Sexually Transmitted Diseases. 2017 June 15;2017:e7075601.

3. Marrazzo JM, del Rio C, Holtgrave DR, Cohen MS, Kalichman SC, Mayer KH, et al. HIV prevention in clinical care settings: 2014 recommendations of the International Antiviral Society-USA Panel. JAMA. 2014 July 23;312(4):390–409.

4. Coffin JM, Hughes SH, Varmus HE. Course of Infection with HIV and SIV. In: Retroviruses [Internet]. Cold Spring Harbor Laboratory Press; 1997 [cited 2025 Oct 12]. Available from: https://www.ncbi.nlm.nih.gov/books/NBK19374/

5. Walker B, McMichael A. The T-Cell Response to HIV. Cold Spring Harb Perspect Med. 2012 Nov;2(11):a007054.

6. Doitsh G, Greene WC. Dissecting How CD4 T Cells Are Lost During HIV Infection. Cell Host Microbe. 2016 Mar 9;19(3):280–91.

7. Oguntibeju OO. Quality of life of people living with HIV and AIDS and antiretroviral therapy. HIV AIDS (Auckl). 2012 Aug 6;4:117–24.

8. HIV and AIDS [Internet]. [cited 2024 Jan 8]. Available from: https://www.who.int/news-room/fact-sheets/detail/hiv-aids

9. Ojikutu BO, Zheng H, Walensky RP, Lu Z, Losina E, Giddy J, et al. Predictors of mortality in patients initiating antiretroviral therapy in Durban, South Africa. S Afr Med J. 2008 Mar;98(3):204–8.

10. Ayele W, Mulugeta A, Desta A, Rabito FA. Treatment outcomes and their determinants in HIV patients on Anti-retroviral Treatment Program in selected health facilities of Kembata and Hadiya zones, Southern Nations, Nationalities and Peoples Region, Ethiopia. BMC Public Health. 2015 Aug 27;15:826.

11. Jerene D, Endale A, Hailu Y, Lindtjørn B. Predictors of early death in a cohort of Ethiopian patients treated with HAART. BMC Infect Dis. 2006 Sept 1;6:136.

12. Brinkhof MWG, Boulle A, Weigel R, Messou E, Mathers C, Orrell C, et al. Mortality of HIV-Infected Patients Starting Antiretroviral Therapy in Sub-Saharan Africa: Comparison with HIV-Unrelated Mortality. PLOS Medicine. 2009 Apr 28;6(4):e1000066.

13. HIV AIDS prevalence rate in Cameroon in 2023∼ Targeting Plus | GPC [Internet]. 2024 [cited 2025 Oct 12]. Available from: https://hivpreventioncoalition.unaids.org/en/news/hiv-aids-prevalence-rate-cameroon-2023-targeting-plus

14. Luma HN, Mboringong F, Doualla MS, Nji M, Donfack OT, Kamdem F, et al. Mortality in Hospitalised HIV/AIDS Patients in a Tertiary Centre in Sub-Saharan Africa: Trends Between 2007 and 2015, Causes and Associated Factors. The Open AIDS Journal [Internet]. 2018 Nov 30 [cited 2024 Jan 9];12(1). Available from: https://openaidsjournal.com/VOLUME/12/PAGE/162/

15. Owachi D, Akatukunda P, Nanyanzi DS, Katwesigye R, Wanyina S, Muddu M, et al. Mortality and associated factors among people living with HIV admitted at a tertiary-care hospital in Uganda: a cross-sectional study. BMC Infectious Diseases. 2024 Feb 22;24(1):239.

16. CMR. Cameroon - Enquête Démographique et de Santé 2018 [Internet]. 2020 [cited 2025 Apr 30]. Available from: https://microdata.worldbank.org/index.php/catalog/3717

17. Sieleunou I, Souleymanou M, Schönenberger AM, Menten J, Boelaert M. Determinants of survival in AIDS patients on antiretroviral therapy in a rural centre in the Far-North Province, Cameroon. Tropical Medicine & International Health. 2009;14(1):36–43.

18. Poka-Mayap V, Pefura-Yone EW, Kengne AP, Kuaban C. Mortality and its determinants among patients infected with HIV-1 on antiretroviral therapy in a referral centre in Yaounde, Cameroon: a retrospective cohort study. BMJ Open. 2013 July 1;3(7):e003210.

19. Kyaw A, Sawangdee Y, Hunchangsith P, Pattaravanich U. Survival Rate and Socio-Demographic Determinants of Mortality in Adult HIV/AIDS Patients on Anti-Retrovial Therapy (ART) in Myanmar: a Registry Based Retrospective Cohort Study 2005-2015. Journal of Health Research [Internet]. 2017 [cited 2025 May 13]; Available from: https://www.semanticscholar.org/paper/Survival-Rate-and-Socio-Demographic-Determinants-of-Kyaw-Sawangdee/666fbc3f825ff79cd1726d780d1d003136f954da

20. Morineau G, Vun MC, Barennes H, Wolf RC, Song N, Prybylski D, et al. Survival and quality of life among HIV-positive people on antiretroviral therapy in Cambodia. AIDS Patient Care STDS. 2009 Aug;23(8):669–77.

21. Patterns and trends in mortality among HIV-infected and HIV-uninfected patients in a major Internal Medicine Unit in Yaoundé, Cameroon: a retrospective cohort study - Josephine Mbuagbaw, Ahmadou M Jingi, Jean Jacques N Noubiap, Arnaud D Kaze, Jobert Richie N Nansseu, Jean Joel R Bigna, Edvine Wawo Yonta, Kathleen Ngu Blackett, 2016 [Internet]. [cited 2025 Oct 12]. Available from: https://journals.sagepub.com/doi/10.1177/2054270416654859?utm_source=chatgpt.com

22. Chimbetete C, Shamu T, Roelens M, Bote S, Mudzviti T, Keiser O. Mortality trends and causes of death among HIV positive patients at Newlands Clinic in Harare, Zimbabwe. PLoS One. 2020 Aug 27;15(8):e0237904.

23. Rimamnunra GN, Utoo PM, Ngwoke K, Bako IA, Akwaras AN, Swende LT, et al. Prevalence of Non-Communicable Diseases among HIV Positive Patients on Antiretroviral Therapy at a Tertiary Health Facility in Makurdi, North-Central, Nigeria. The Nigerian Health Journal. 2023 Oct 23;23(3):734–40.

24. Isa SE, Oche AO, Kang’ombe AR, Okopi JA, Idoko JA, Cuevas LE, et al. Human Immunodeficiency Virus and Risk of Type 2 Diabetes in a Large Adult Cohort in Jos, Nigeria. Clin Infect Dis. 2016 Sept 15;63(6):830–5.

25. Adeoti AO, Dada M, Elebiyo T, Fadare J, Ojo O. Survey of antiretroviral therapy adherence and predictors of poor adherence among HIV patients in a tertiary institution in Nigeria. The Pan African Medical Journal [Internet]. 2019 July 31 [cited 2025 Oct 12];33(277). Available from: https://www.panafrican-med-journal.com//content/article/33/277/full

26. Sharma A, Hoover DR, Shi Q, Gustafson D, Plankey MW, Hershow RC, et al. Relationship between Body Mass Index and Mortality in HIV-Infected HAART Users in the Women’s Interagency HIV Study. PLoS One. 2015;10(12):e0143740.

27. Tadesse K, Haile F, Hiruy N. Predictors of Mortality among Patients Enrolled on Antiretroviral Therapy in Aksum Hospital, Northern Ethiopia: A Retrospective Cohort Study. PLoS One. 2014 Jan 31;9(1):e87392.

